# The prediction of cardiovascular events by phenotype of central and peripheral blood pressure in subjects without hypertension

**DOI:** 10.1101/2023.02.03.23285459

**Authors:** Min-Sik Kim, Gee-Hee Kim

## Abstract

**Background:** Hypertension (HBP) is a common disease associated with aging, but the rate of recognition and control of the condition remains low. Most guidelines related to HBP have consisted of only peripheral blood pressure (BP) measurement. However, according to many studies, central BP (CBP) has a clearer relationship with the prediction of cardiovascular (CV) events than does peripheral BP and can more clearly express an individual’s BP status. Therefore, we aimed to evaluate the effect of CBP on the prediction of CV events and to investigate the prediction of CV events by phenotype of central and peripheral BP in subjects without hypertension.

**Method:** A total of 2,910 patients were enrolled from June 2011 to December 2016 and were followed up through October 2022. CBP was measured using radial tonometry. The primary endpoint was a composite outcome.

**Result:** The median follow-up period for enrolled patients was 7.5 years. Out of a total of 722 patients (mean age of 52.5 ± 13.7 years) without HBP, 21 patients (2.9%) had events of the primary endpoint during the follow-up period. Systolic BP averaged 126 mmHg (±15 mmHg) in the event-free group and 136 mmHg (±15 mmHg) in the CV event group, while CBP measured 115 mmHg (±16 mmHg) in the event-free group and 126 mmHg (±16 mmHg) in the CV event group. In a Cox proportional hazards model, every 10 mmHg increase in CBP and systolic BP showed an increase in risk of 30% and 40%, respectively. Isolated central systolic hypertension and dual central and peripheral systolic hypertension showed 4.9% and 6% of the CV event rate, respectively (p=0.897).

**Conclusion:** Irrespective of the brachial BP status, isolated central hypertension increased CV events. Therefore, to prevent CV events, it is essential to control not only peripheral BP but also CBP.

## Introduction

High blood pressure (BP) has doubled worldwide since 1990, according to a report based on large data [1]. In addition, the importance of high BP is continuously emphasized because controlling BP with low cost is effective in preventing cerebrovascular accidents as well as vascular diseases such as ischemic heart disease and kidney disease [2]. The report also emphasized that the diagnosis rate or treatment rate is lower than the increasing prevalence rate. According to a fact sheet released in Korea in 2022, it is higher than the global average, but about 30% does not recognize hypertension (HBP), and the treatment rate tends to decrease [3]. There are several hurdles and limitations contributing to this. Depending on a number of factors, an individual’s BP status can change significantly [4]. However, office BP has limitations in evaluating such BP variability. It has recently been emphasized that BP variability is more important for targeting organ damage [5]. Even if a doctor tries to proceed with 24-hour ambulatory BP monitoring (ABPM), there are hurdles in examination cooperation, such as lack of patient insight.

One of the out-of-office BP measurements, central blood pressure (CBP), has long been considered a better predictor than brachial BP for CV event risk, and meta-analyses have been sufficiently conducted accordingly [6]. In addition, the non-invasive central hemodynamics measurement method was sufficiently validated [7, 8]. Given the anatomical proximity of the heart, brain, and kidney to the central arteries and the role of arterial stiffness in cardiovascular disease, CBP is thought to be more closely related than brachial BP. In addition, there have been reports that CBP may be different due to the characteristics of blood vessels even in people with the same brachial BP [9]. In recent meta-analysis, patients with increased CBP but normotensive peripheral BP had increased CV events [10]. However, CBP has not yet been included in the guidelines for BP management. Since the SPRINT trial emphasized the advantageous aspect of tight BP control [11], it may be necessary to focus on finding patients who need more aggressive management that can be overlooked only by office BP measurement. In addition, it is still controversial whether assessment of CBP is necessary even in the presence of normal brachial BP.

Therefore, in this study, we evaluate the role of CBP by measuring brachial systolic BP (SBP) and CBP in patients who have never been treated for HBP before. Also, we investigate the prediction of CV events by phenotype of central and peripheral BP in subjects without hypertension.

## Method

### Study Population

Patients were enrolled from July 2011 to December 2016, and blood pressure was measured non-invasively using the same device as in our previous study. Among a total of 2910 patients, the following patients were excluded: those who had a CV event within the previous 3 months; those who were previously diagnosed with hypertension; or those who were taking blood pressure medications including ACEi/ARB, CCB, beta blockers, and diuretics. Patients with irregular rhythms or those whose brachial blood pressure could not be measured due to brachial artery stenosis were also excluded from the study. Thus, a total of 720 patients were analyzed.

There was no industry involvement in the design, implementation, or data analysis of this study. The present study was a single-center retrospective study and was approved by the Institutional Review Board of St. Vincent’s Hospital (VC22RISI0070).

### Blood pressure measurement

All measurements of hemodynamic indices were taken as the average of the values obtained by measuring twice after sitting on a chair in a quiet room with a constant temperature and resting for more than 5 minutes. For measurement, a cuff ocillometric device was used in the upper right arm, and at the same time, the waveform of the left radial artery was automatically measured through a multi-element tonometry sensor composed of 40 micro-transducers; the measurement was sufficiently taken for more than 30 seconds until a stable waveform was obtained. The signal was digitized 500 Hz, and the hold-down pressure of the sensor unit was automatically adjusted for each participant. The method used to measure CBP was the same as in a previous study [12].

### Clinical and biochemical assessments

Considering the effect of circadian variations, blood sampling was conducted between 8:00 and 9:30 am after fasting for 12 to 14 hours. Total cholesterol levels followed standard enzymatic methods, HDL was measured after precipitating VLDL, and LDL was calculated according to the Friedewald formula. eGFR was calculated using the CKD-EPI formula. It was the same condition as in our previous studies [12].

### Outcome

The primary endpoint was a composite outcome of CV events, including chronic coronary syndrome, acute coronary syndrome, heart failure (elevation of NT-proBNP), stroke, TIA, and PAD with ABI<0.9. During the follow-up period, a cardiologist reviewed the patient medical records to determine whether or not a primary endpoint occurred.

### Statistical analysis

Continuous variables were expressed as ± standard deviation of the mean, and categorical variables were expressed as absolute and relative frequencies. T tests were used to compare means between two groups, and ratios were tested for both tables and chi-square. To determine the independent predictors of the primary endpoint, multivariate analysis using Cox proportional hazards regression models was used for significant risk factors found to be important and primary endpoints in univariate analysis. Multivariate analysis was presented using constrained cubic spline curves. All statistical analyses were performed on version 3.6.3.

## Results

The median follow-up period for enrolled patients was 7.5 years, with an average age of 52.5 ± 13.7 years. Of the 720 patients in the study group, 47.5% were men and 52.5% were women. The average body mass index (BMI) was 23.75 ± 3.2 kg/m^2^, and 21 patients (2.9%) had primary endpoint events during the follow-up period. One of them died of cardiac death, 9 patients were diagnosed with vascular disease including ACS and coronary reperfusion, and 6 experienced stroke, TIA, or brain hemorrhage. Heart failure occurred in 5 patients.

Table 1 shows the baseline characteristics of the participants by classifying them into patients with and without a primary endpoint event. There was a statistically significant difference between the two groups only in age. In addition, CBP, SBP, pulse pressure (PP), and central PP (CPP) were statistically significantly different between the two groups.

**Table 1.** Baseline Characteristics of the Participants.

| Variables | CV event (-)<br>699 | CV event (+)<br>21 | P-value |
| --- | --- | --- | --- |
| Age, years (SD) | 52.17 (13.63) | 64.48 (10.41) | <0.001 |
| BMI, kg/m<sup>2</sup> (SD) | 23.69 (3.20) | 23.33 (3.00) | 0.610 |
| Smoking, N (%) | 164 (24.50) | 9 (45.00) | 0.069 |
| Diabetes, N (%) | 107 (15.30) | 7 (33.30) | 0.054 |
| HbA1c, % (SD) | 7.34 (2.35) | 6.33 (0.23) | 0.460 |
| eGFR, mL/min/1.73m<sup>2</sup> (SD) | 101.43 (16.92) | 93.82 (24.99) | 0.065 |
| TC, mg/dl (SD) | 188.64 (40.47) | 182.81 (45.14) | 0.572 |
| TG, mg/dl (SD) | 125.11 (94.11) | 128.75 (83.08) | 0.879 |
| LDL, mg/dl (SD) | 114.72 (34.51) | 106.57 (29.62) | 0.383 |
| HDL, mg/dl (SD) | 45.98 (12.29) | 41.50 (12.39) | 0.180 |
| Statin use (SD) | 107 (15.30) | 4 (19.00) | 0.872 |
| CBP, mmHg (SD) | 115.69 (16.43) | 125.86 (15.26) | 0.005 |
| SBP, mmHg (SD) | 125.86 (15.37) | 136.38 (15.87) | 0.002 |
| DBP, mmHg (SD) | 76.55 (10.54) | 78.05 (12.54) | 0.524 |
| PP, mmHg (SD) | 49.30 (11.34) | 58.33 (12.63) | <0.001 |
| CPP, mmHg (SD) | 39.24 (12.19) | 47.35 (10.57) | 0.003 |
| Alx@75, (SD) | 79.24 (14.40) | 82.71 (11.43) | 0.274 |
| Heart Rate, (SD) | 71.78 (12.53) | 72.14 (12.02) | 0.897 |

**Abbreviation:** Alx@75, Augmentation index adjusted with a heart rate of 75; BMI, Body mass index; CI, Confidential interval; eGFR, estimated Glomerular filtration rate; TC, Total cholesterol; TG, Triglyceride; LDL, Low density lipoprotein; HDL, high density lipoprotein; CBP, Central blood pressure; SBP, Systolic blood pressure; DBP, Diastolic blood pressure; PP, Pulse pressure;

Table 2 presents the results of univariate Cox proportional hazards ratio models for each variable. Table 3 shows a multivariable Cox proportional hazards model adjusted for age and smoking. After adjustment, statistically significant increases in risk of 30% for CBP and 40% for SBP were confirmed, as was an increase in risk of about 40% for PP, but CPP did not acquire statistical significance.

**Table 2.**
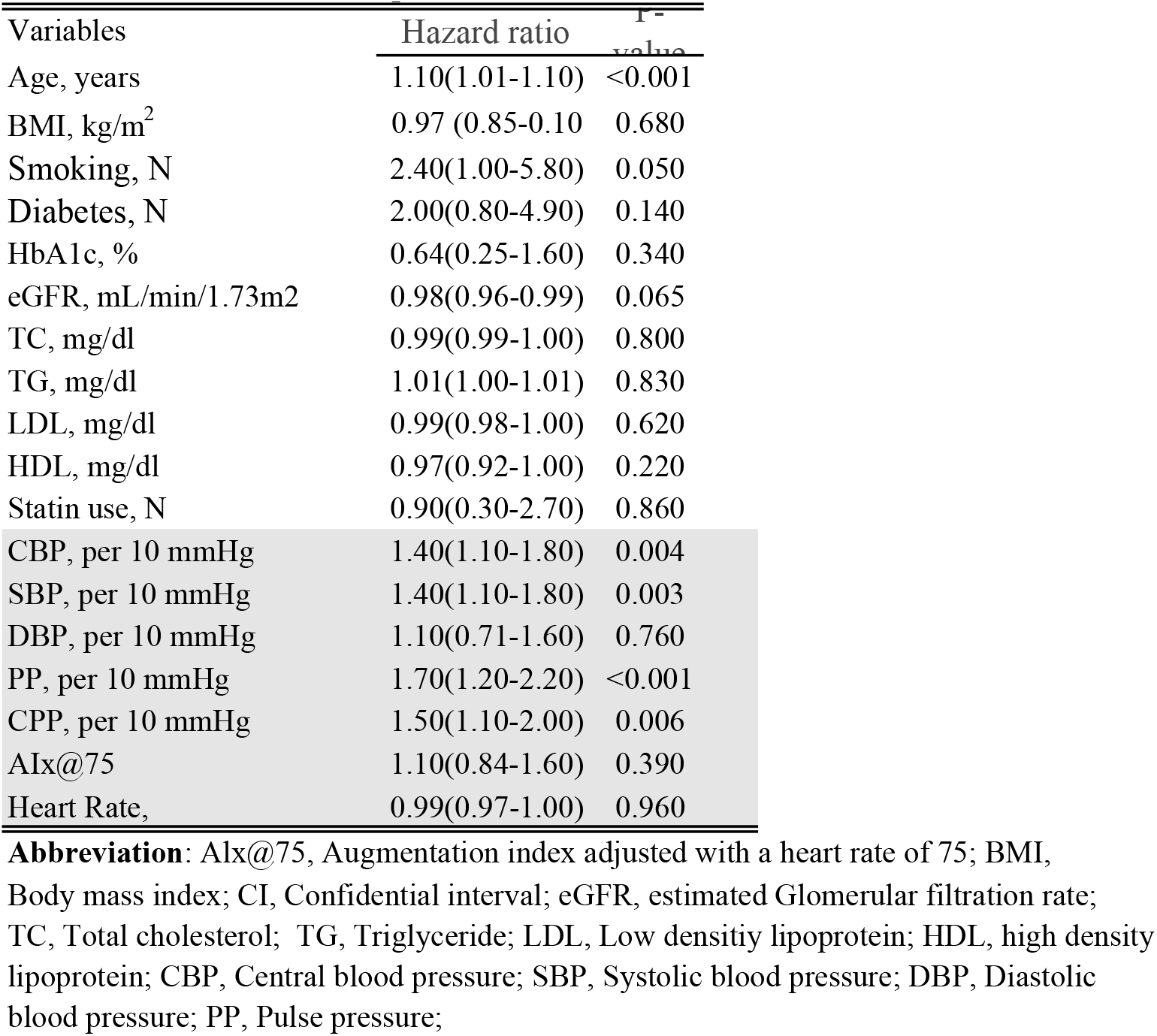
Univariate Cox Proportional Hazards Model

| Variables | Hazard ratio | P-value |
| --- | --- | --- |
| Age, years | 1.10(1.01-1.10) | <0.001 |
| BMI, kg/m<sup>2</sup> | 0.97 (0.85-0.10) | 0.680 |
| Smoking, N | 2.40(1.00-5.80) | 0.050 |
| Diabetes, N | 2.00(0.80-4.90) | 0.140 |
| HbA1c, % | 0.64(0.25-1.60) | 0.340 |
| eGFR, mL/min/1.73m<sup>2</sup> | 0.98(0.96-0.99) | 0.065 |
| TC, mg/dl | 0.99(0.99-1.00) | 0.800 |
| TG, mg/dl | 1.01(1.00-1.01) | 0.830 |
| LDL, mg/dl | 0.99(0.98-1.00) | 0.620 |
| HDL, mg/dl | 0.97(0.92-1.00) | 0.220 |
| Statin use, N | 0.90(0.30-2.70) | 0.860 |
| CBP, per 10 mmHg | 1.40(1.10-1.80) | 0.004 |
| SBP, per 10 mmHg | 1.40(1.10-1.80) | 0.003 |
| DBP, per 10 mmHg | 1.10(0.71-1.60) | 0.760 |
| PP, per 10 mmHg | 1.70(1.20-2.20) | <0.001 |
| CPP, per 10 mmHg | 1.50(1.10-2.00) | 0.006 |
| Alx@75 | 1.10(0.84-1.60) | 0.390 |
| Heart Rate, | 0.99(0.97-1.00) | 0.960 |

**Abbreviation:** Alx@75, Augmentation index adjusted with a heart rate of 75; BMI, Body mass index; CI, Confidential interval; eGFR, estimated Glomerular filtration rate; TC, Total cholesterol; TG, Triglyceride; LDL, Low density lipoprotein; HDL, high density lipoprotein; CBP, Central blood pressure; SBP, Systolic blood pressure; DBP, Diastolic blood pressure; PP, Pulse pressure;

**Table 3.** Multivariate Cox Proportional Hazards Model

| Variables | Hazard ratio<br>(95% CI) | P-value |
| --- | --- | --- |
| CBP, per 10 mmHg | 1.30(1.01-1.70) | 0.021 |
| SBP, per 10 mmHg | 1.40(1.10-1.80) | 0.014 |
| DBP, per 10 mmHg | 1.40(0.91-2.00) | 0.130 |
| PP, per 10 mmHg | 1.40(1.01-2.10) | 0.042 |
| CPP, per 10 mmHg | 1.20(0.87-1.80) | 0.220 |
| Alx@75 | 0.99(0.70-1.50) | 0.840 |
| Heart Rate | 0.99(0.98-1.00) | 0.590 |

**Abbreviation:** Alx@75, Augmentation index adjusted with a heart rate of 75; CBP, Central interval; SBP, Systolic blood pressure; DBP, Diastolic blood pressure; PP, Pulse pressure

Figure 1 shows a restricted cubic spline curve for the risk of CV event occurrence according to the increase in CBP in subjects without HBP. In this analysis, the point considered to be the optimal CBP was 110 mmHg. In addition, the risk of CV event occurrence according to the increase in BP showed a J-curve pattern.

**Figure 1.**
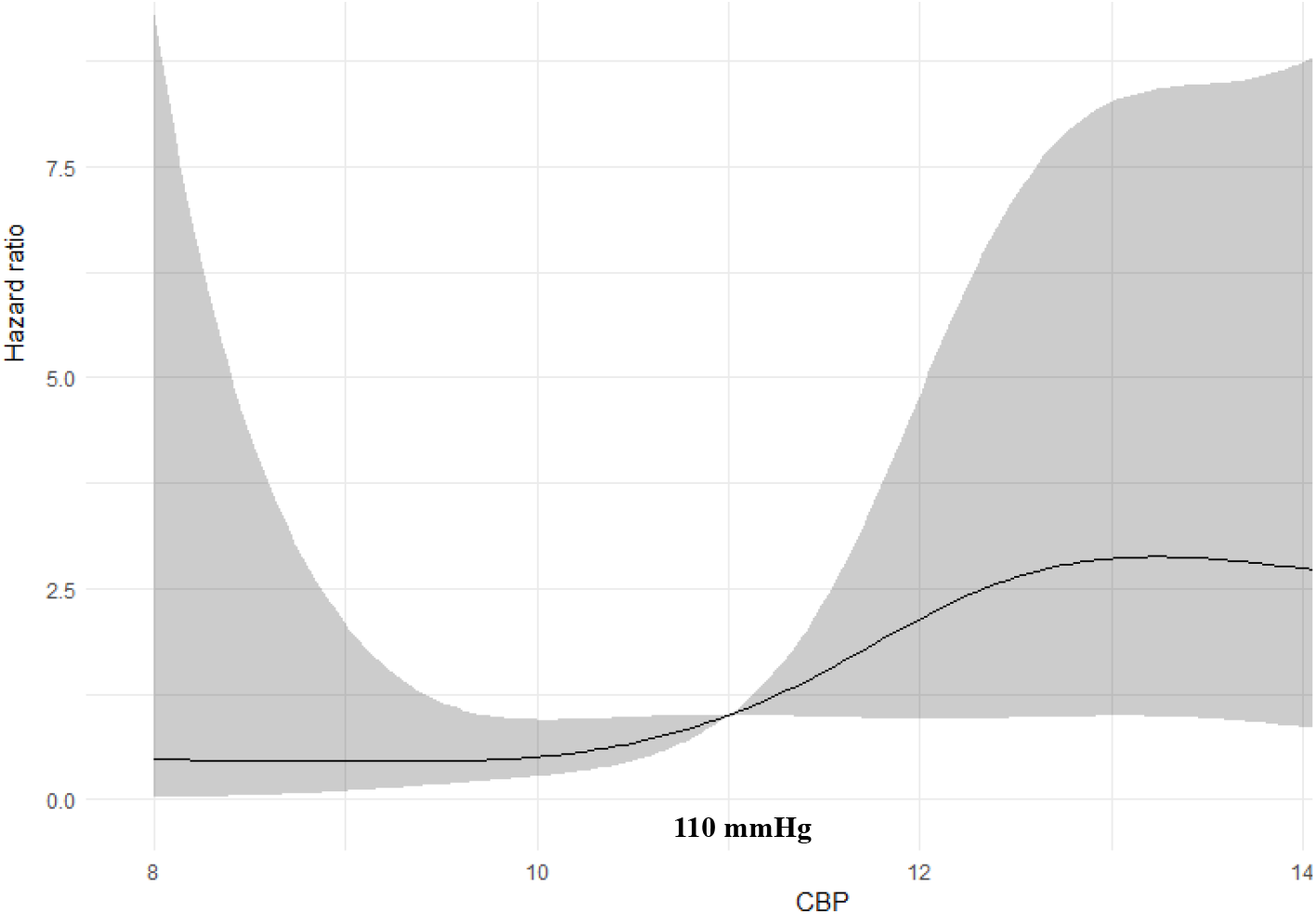
Restricted cubic spline curve of CBP; the lowest CBP of cumulative incidence of the primary outcome was 110 mmHg.

In Figure 2, using a categorical approach, CBP at 120mmHg and SBP at 140mmHg might be divided into normotensive and hypertensive levels based on our previous study and Korea guideline [3, 13-15]. Group 1 (isolated central hypertension group, consisting of high CBP and normal SBP) had a total of 162 patients, with average values for age, BMI, CBP, SBP, DBP, PP, and CPP of 57.2 ± 10.8 years, 24.2 ± 3.1 kg/m^2^, 126.4 ± 4.5 mmHg, 132.1 ± 4.8 mmHg, 81.0 ± 7.9 mmHg, 51.0 ± 8.0 mmHg, and 45.5 ± 8.6 mmHg, respectively. There were 116 people in Group 2 (both systolic hypertension group, consisting of high SBP and high CBP). The average values for age, BMI, CBP, SBP, DBP, PP, and CPP were 58.0 ± 12.0 years, 24.1 ± 3.18 kg/m^2^, 140.8 ± 11.5 mmHg, 150.3 ± 9.1 mmHg, 87.1 ± 11.3 mmHg, 63.2 ± 12.6 mmHg, and 53.6 ± 14.1 mmHg, respectively. However, there was no difference between the two groups in baseline characteristics. Isolated central systolic hypertension and both systolic hypertension showed 4.9% and 6% of the CV event rate, respectively. There was no difference between the two groups (p=0.897). Group 3 (consisting of normal SBP and CBP) had a total of 432 patients, and the average values for age, BMI, CBP, SBP, DBP, PP, and CPP were 49.6 ± 14.0 years, 23.3 ± 3.2 kg/m^2^, 105.5 ± 9.6 mmHg, 117.0 ± 10.2 mmHg, 72.0 ± 8.4 mmHg, 45.0 ± 8.5 mmHg, and 33.5 ± 7.9 mmHg, respectively. Group 4 (normal CBP and high SBP) consisted of a small number of 10 people.

**Figure 2.**
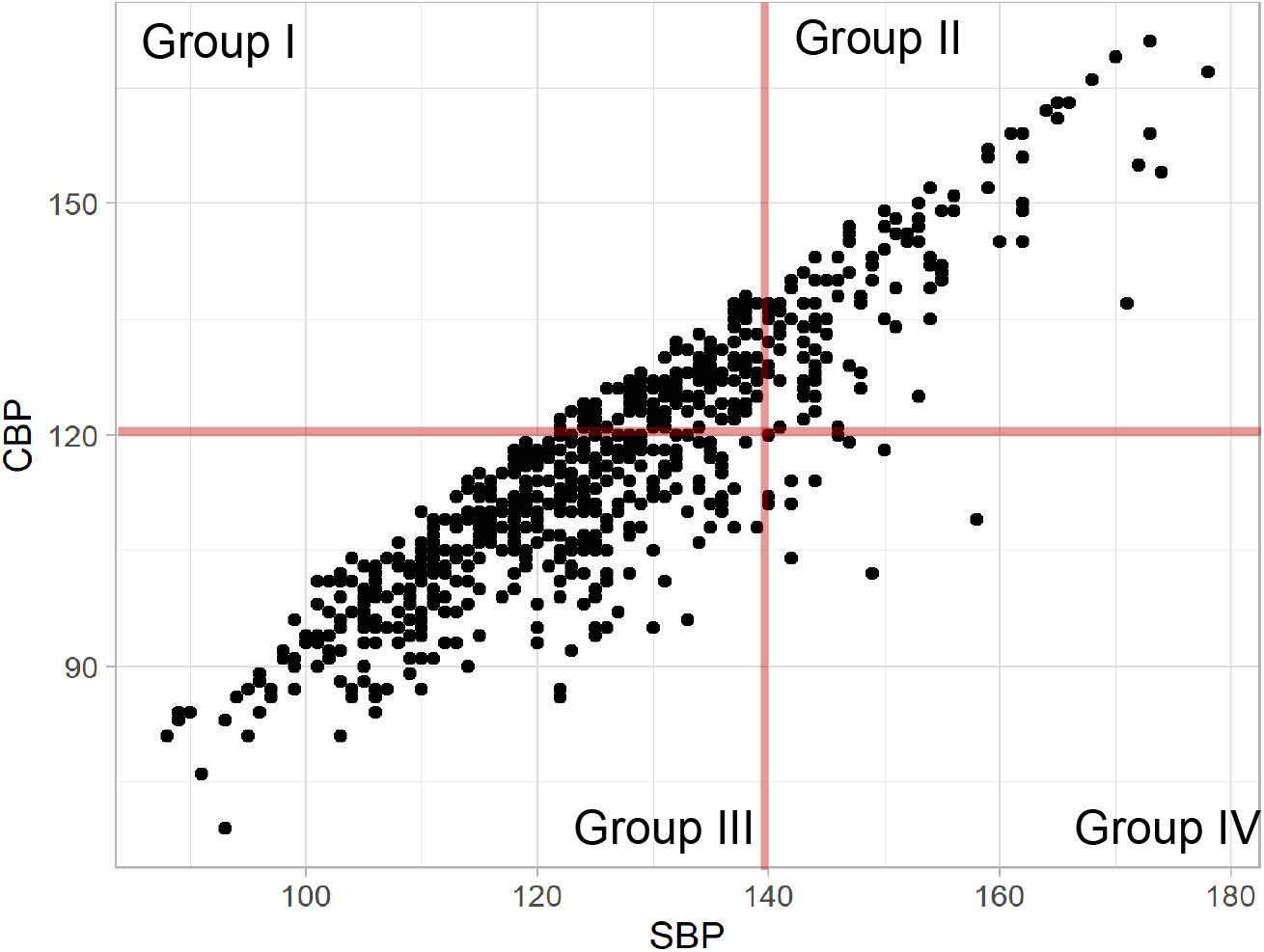
CBP corresponding to each SBP is represented as a dot; SBP and CBP are in a direct proportional relationship

## Discussion

This study found that CVD risk according to CBP in patients without HBP had a J-shaped curve, and the lowest level of CBP was 110 mmHg. Isolated central hypertension and both systolic hypertension showed 4.9% and 6% of the CV event rate, respectively.

Generally, those who have never been diagnosed with HBP and have never taken antihypertensive drugs may have fewer CV events than hypertensive patients. According to a fact sheet released in Korea in 2022 [3], about 30% does not recognize HBP. In our study, 17.5% of those without recognizing high BP diagnosed as hypertension when 140 mmHg was the cut-off value of peripheral BP. In addition, it was investigated that CV events occurred in 6.3%, so it seems that not a few events occurred.

In clinical practice, BP is routinely measured at the brachial artery. However, follow-up in hypertensive patients by also measuring CBP can be more advantageous as a predictor of CV disease, as seen in the Café study [16]. In addition, it is thought that CBP may be more advantageous than brachial BP, which is mainly measured at the office, in evaluating target organ damage in normotensive patients [17, 18]. Therefore, our previous study showed that CV risk increased rapidly at a CBP of 120 mmHg or higher through previously studied CBP data, irrespective of the brachial BP status [13-15]. In a recent meta-analysis, optimal CBP and central hypertension were defined as less than 110 mmHg and 120 mmHg, respectively, based on the primary CV endpoint with a 5-year probability of cohort data [10]. Similar to the previous study, the lowest CBP was 110 mmHg among the cases of CV events in our study. In addition, the threshold of central hypertension was categorized at 120 mmHg, and in this case, the incidence of CV events tended to increase regardless of brachial SBP.

In our study, the CV event rate was 6% in patients with both systolic hypertension who needed conventional hypertensive treatment, whereas the CV event rate in patients with isolated central hypertension, who were not subject to aggressive hypertension treatment, was 4.9%. In the previous study, the cumulative CV events increased gradually from concordant central and brachial normotensive over isolated brachial hypertension to isolated central hypertension and onward to concordant central and brachial hypertension with no difference between isolated central hypertension and concordant hypertension, although 130 mmHg was the cut-off value for peripheral BP. Based on these results, isolated central hypertension should not be overlooked even if brachial BP is normal [17, 19].

A study found that most patients with isolated systolic hypertension (ISH) and normal CBP had white coat hypertension (WCH) [20] [21], so these patients meet the diagnostic requirements for WCH and may require ABPM. However, if the ABPM test is not available or if a young patient with a high BMI has high BP, evaluation with CBP may be necessary rather than using many drugs. The tendency of CBP to be inconsistent with brachial SBP is probably because central hemodynamic measures show a closer correlation with vascular aging, and vascular aging-associated hypertension has been reported in Asia. Based on this, it is thought that this tendency was also reflected in this study [22, 23].

In our study, there is a limitation that ABPM was not performed. It is thought that more information would have been obtained by classifying patients with masked hypertension and WCH through ABPM [24-26] and analyzing their CBP patterns. This is because our study is a retrospective observational study, and the number of participants was insufficient because we excluded those who were diagnosed with hypertension or who were taking antihypertensive medications for other reasons. In addition, aggressive evaluation was not conducted on participants who met these criteria. In future studies, survival analysis should be considered with a longer study period and a larger number of participants.

## Conclusion

In this study, irrespective of brachial BP status, isolated central hypertension increased CV events. In addition, CBP not only acts as a predictor in CV outcomes, but is also thought to play a role in risk stratification of patients, which may be insufficient with brachial BP measurement. Therefore, to prevent CV events, it is essential to control not only peripheral BP but also CBP.

## Data Availability

The datasets used and/or analyzed during the current study are available from the corresponding author upon reasonable request.

